# Appealing for Efficient, Well Organized Clinical Trials on COVID-19

**DOI:** 10.1101/2020.03.05.20031476

**Authors:** Yang Zhao, Yongyue Wei, Sipeng Shen, Mingzhi Zhang, Feng Chen

## Abstract

The rapid emergence of clinical trials on COVID-19 stimulated a wave of discussion in scientific community. We reviewed the characteristics of interventional trials from Chinese Clinical Trial Registration (ChiCTR) and ClinicalTrials.gov. A total of 171 COVID-19-related interventional trials were identified on Feb 22nd, 2020. These trials are classified into 4 categories based on treatment modalities, including chemical drugs, biological therapies, traditional Chinese medicine treatments and other therapies. Our analysis focused on the issues of stage, design, randomization, blinding, primary endpoints definition and sample size of these trials. We found some studies with potential defects including unreasonable design, inappropriate primary endpoint definition, insufficient sample size and ethical issue. Clinical trials on COVID-19 should be designed based on scientific rules, ethics and benefits for patients.

## Introduction

As of February 29, 2020, approximately 85,000 cases of coronavirus disease 2019 (COVID-19) have been confirmed worldwide, with nearly 3,000 deaths occured. Recently, many COVID-19-related interventional clinical trials have emerged in China, the original and high-incidence area of COVID-19^1^. The rapid emergence of these trials stimulated a wave of discussion. Herein, we reviewed the characteristics of interventional trials from Chinese Clinical Trial Registration (ChiCTR) and ClinicalTrials.gov.^2,3^

## Methods

The data of interventional trials from ChiCTR and ClinicalTrials.gov were retrieved updated on February 22, 2020. Two authors (Zhao and Shen) were independently responsible for collecting the relevant information, including clinical phase, study design, presence or absence of randomization, blinding, sample size, severity of disease and source of samples.

## Results

A total of 171 COVID-19-related interventional trials were identified (138 from ChiCTR and 33 from ClinicalTrials.gov). The registration date distribution of these trials was shown in Figure 1. Of the trials registered at ChiCTR, 120 were approved by the institutional review boards. The 171 trials are classified into 4 categories based on treatment modalities, including chemical drugs (CDs), biological therapies (BTs), traditional Chinese medicine (TCM) treatments and other therapies. The characteristics of these trials were summarized in Table 1.

**Table 1.** Characteristics of the 171 Interventional Clinical Trials [n(%)]

|  | Chemical drugs*<br>(n=61) | Biological therapies*<br>(n=42) | TCM treatment*<br>(n=59) | Other therapies<br>(n=9) | Total<br>(n=171) |
| --- | --- | --- | --- | --- | --- |
| Phase |  |  |  |  |  |
| I and II | 20 (32.8) | 27 (64.3) | 28 (47.5) | 6 (66.7) | 81 (47.4) |
| II/III and III | 8 (13.1) | 2 (4.7) | 1 (1.7) | 0 (0) | 11 (6.4) |
| IV | 22 (36.1) | 7 (16.7) | 16 (27.1) | 0 (0) | 45 (26.3) |
| Others | 11 (18.0) | 6 (14.3) | 14 (23.7) | 3 (33.3) | 34 (19.9) |
| Design |  |  |  |  |  |
| Single-arm | 5 (8.2) | 8 (19.0) | 4 (6.8) | 0 (0) | 17 (9.9) |
| Parallel-arm | 56 (91.8) | 34 (81.0) | 54 (91.5) | 7 (77.8) | 151 (88.3) |
| Factorial design** | 0 (0.0) | 0 (0.0) | 1 (1.7) | 2 (22.2) | 3 (1.8) |
| Randomization |  |  |  |  |  |
| No | 4 (6.6) | 4 (9.5) | 9 (15.2) | 0 (0) | 17 (9.9) |
| Yes | 50 (82.0) | 28 (66.7) | 46 (78.0) | 8 (88.9) | 132 (77.2) |
| Not applicable | 7 (11.4) | 10 (23.8) | 4 (6.8) | 1 (11.1) | 22 (12.9) |
| Blinding |  |  |  |  |  |
| Open | 23 (37.7) | 16 (38.0) | 21 (35.6) | 3 (33.3) | 63 (36.9) |
| Single | 5 (8.2) | 2 (4.8) | 1 (1.7) | 0 (0) | 8 (4.7) |
| Double/triple/quadruple | 10 (16.4) | 2 (4.8) | 5 (8.5) | 0 (0) | 17 (9.9) |
| Unavailable | 14 (23.0) | 11 (26.2) | 23 (39.0) | 4 (44.4) | 52 (30.4) |
| Not applicable | 9 (14.7) | 11 (26.2) | 9 (15.2) | 2 (22.2) | 31 (18.1) |
| Sample size |  |  |  |  |  |
| ≤50 | 15 (24.6) | 17 (40.5) | 3 (5.1) | 1 (11.2) | 36 (21.1) |
| 50-100 | 21 (34.4) | 13 (31.0) | 20 (33.9) | 3 (33.3) | 57 (33.3) |
| 100-400 | 23 (37.7) | 10 (23.8) | 31 (52.5) | 3 (33.3) | 67 (39.2) |
| >400 | 2 (3.3) | 2 (4.7) | 5 (8.5) | 2 (22.2) | 11 (6.4) |
| Number of primary endpoints |  |  |  |  |  |
| 1 | 37 (60.6) | 25 (59.5) | 25 (42.4) | 1 (11.1) | 88 (51.5) |
| 2 | 12 (19.7) | 5 (11.9) | 11 (18.6) | 4 (44.4) | 32 (18.7) |
| ≥3 | 12 (19.7) | 12 (28.6) | 23 (39.0) | 4 (44.4) | 51 (29.8) |
| Type of primary endpoint<sup>#</sup> |  |  |  |  |  |
| Nucleic acid amplification testing | 25 (41.0) | 10 (23.8) | 13 (22.0) | 2 (22.2) | 50 (29.2) |
| Fatality | 11 (18.0) | 4 (9.5) | 5 (8.5) | 0 (0) | 20 (11.7) |
| Clinical symptoms | 39 (63.9) | 28 (66.7) | 51 (86.4) | 7 (77.8) | 125 (73.1) |
| Imaging | 13 (21.3) | 8 (19.0) | 11 (18.6) | 0 (0) | 32 (18.7) |
| Lung function | 14 (23.0) | 12 (28.6) | 17 (28.8) | 4 (44.4) | 47 (27.5) |
| TCM symptoms | 0 (0) | 0 (0) | 7 (11.9) | 0 (0) | 7 (4.1) |
| Immunology | 4 (6.6) | 5 (11.9) | 4 (6.8) | 1 (11.1) | 14 (8.2) |
| Severity of pneumonia<sup>#</sup> |  |  |  |  |  |
| Mild | 46 (75.4) | 27 (64.3) | 36 (61.0) | 4 (44.4) | 64 (66.1) |
| Severe | 19 (31.1) | 32 (76.2) | 17 (28.8) | 2 (22.2) | 70 (40.9) |
| Critically ill | 7 (11.5) | 11 (26.2) | 9 (15.3) | 1 (11.1) | 28 (16.4) |
| Samples***<sup>#</sup> |  |  |  |  |  |
| No | 1 (2.3) | 2 (6.5) | 6 (10.9) | 4 (44.4) | 13 (9.4) |
| Blood | 28 (65.1) | 25 (80.6) | 44 (80.0) | 3 (33.3) | 100 (72.4) |
| Nasopharyngeal swabs | 11 (25.6) | 3 (9.7) | 3 (5.5) | 2 (22.2) | 19 (13.8) |
| Respiratory secretions | 5 (11.6) | 1 (3.2) | 2 (3.6) | 0 (0) | 8 (5.8) |
| Bronchoalveolar lavage Fluid | 2 (4.7) | 0 (0) | 0 (0) | 0 (0) | 2 (1.4) |
| Stool | 1 (2.3) | 0 (0) | 0 (0) | 0 (0) | 1 (0.7) |

**Notes:**
\*Chemical drugs include remdesivir, ritonavir, triavevirin, leflumite, glucocorticoid, hydroxychloroquine, darunavir and cobicistat, etc. Biological therapies include PD1, bevacizumab, natural killer cell therapy and convalescent plasma therapy. TCM treatments include TCM and physical treatment (Qigong and Taiji).
\*\*Three trials are registered as “factorial design”, one for TCM, one for psychological intervention to doctors and nurses, and one for probiotics. However, unlike the general purpose of factorial design, none of the 3 trials aim to find the best “combination” of the treatments.
\*\*\*Trials registered at ClinicalTrials do not provide information about the collected samples.
#Percentages for PE type are calculated by the number of trials using this PE divided by the number of corresponding type of trials. Percentages of severity and samples are calculated in the same manner.

**Figure 1.**
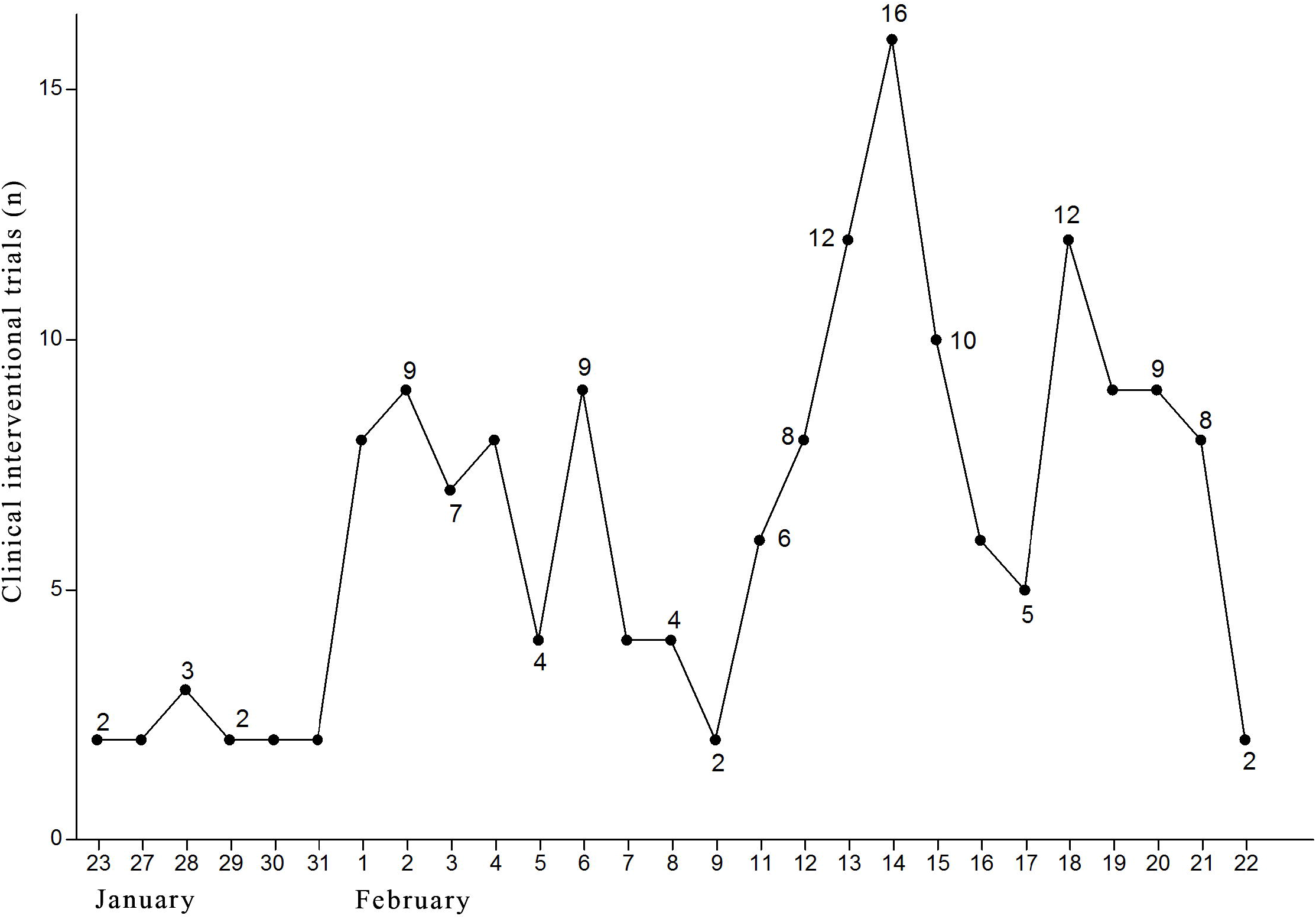
The Registration Date Distribution of the Interventional Clinical Trials

There are 11 confirmatory Phase III or Phase II/III studies (6.4%), 8 of which are for CDs. Most trials (88.3%) use parallel-arm design. Randomization is applied in 132 trials (77.2%), while blinding in only 25 trials (14.6%). The sample sizes of 45.6% trials are more than 100. Two trials on TCM treatments are planned to recruit 800 or more patients. Severe and critically ill patients are respectively recruited in 70 (40.9%) and 28 (16.4%) trials. More than half of the trials (72.4%) are planned to collect blood samples, while only 19 trials (13.8%) to collect nasopharyngeal swabs. More trials on TCM treatment use 2 or more primary endpoints (PEs) than those on CDs or BTs (57.6%, 39.4% and 40.5%, respectively). Some trials even have 6 or more PEs. The most 3 common used PEs are associated with clinical symptoms(73.1%), pathogen(29.2%) and lung function(27.5%). Trials on CDs have greater proportions of pathogen (41.0%) or fatality (18.0%) related PEs than the others. It is worth noting that these trials have registered over 300 distinct PEs totally.

## Discussion

During the outbreak, we are in urgent need of an effective treatment strategies. Through analyzing currently-registered interventional trials, we found some studies with potential defects including unreasonable design, inappropriate PE definition, small sample size and ethical issue.

Determining the appropriate PEs is crucial to address the primary scientific question of the study. The PEs should be defined clearly and specifically to improve the operation quality. Objective endpoints, such as pathogen or imaging results, are preferred to avoid the bias during evaluation.

It is possible that some trials lack statistical power. Assuming a two-side significance level of 0.05 and power of 80%, a trial on mild pneumonia patients should recruit nearly 1,000 patients if the effective rate increases from 90% to 95% for the new treatment. More patients are needed for the trials planning to decrease the fatality of critically ill patients.

Some drugs and therapies under study are short of previous theoretical support for COVID-19. These drugs and therapies may be not beneficial to the patients, leading to non-compliance to ethical standards. These unnecessary trials may also waste medical resources, as well as diverting patients resources. WHO has suggested an establishment of a centralized research program to ensure the most promising researches^4^.

As appealed by some Chinese scientists recently, clinical trials on COVID-19 should be designed based on scientific rules (appropriate controls, randomization, blinding, and sufficient sample size),ethics and patients’ benefits^5^. The standards for clinical trials should not be compromised in such a special time.

## Data Availability

All information used in this article can be retrieved directly from CHiCTR and CLinicalTrials.

## Conflict of Interest Disclosures

None.

## Acknowledgment

This study was funded by the National Natural Science Foundation of China (Grant N o. 81872709) and Key Project of the Natural Science Foundation of the Jiangsu Higher Education Institutions of China (Grant No. 18KJA110004). The authors are grateful to Miss Yuanping Yue and Mrs. Yue Wang for their help on preparing the manuscript.

## Notes

### Competing Interest Statement

The authors have declared no competing interest.

### Funding Statement

This study was funded by the National Natural Science Foundation of China (Grant No. 81872709) and Key Project of the Natural Science Foundation of the Jiangsu Higher Education Institutions of China (Grant No. 18KJA110004). The authors are grateful to Mr. Mingzhi Zhang, Miss Yuanping Yue and Mrs. Yue Wang for their help on preparing the manuscript.

